# Performance of intensive care unit severity scoring systems across different ethnicities

**DOI:** 10.1101/2021.01.19.21249222

**Authors:** Rahuldeb Sarkar, Christopher Martin, Heather Mattie, Judy Wawira Gichoya, David J. Stone, Leo Anthony Celi

## Abstract

**Background:** Despite wide utilisation of severity scoring systems for case-mix determination and benchmarking in the intensive care unit, the possibility of scoring bias across ethnicities has not been examined. Recent guidelines on the use of illness severity scores to inform triage decisions for allocation of scarce resources such as mechanical ventilation during the current COVID-19 pandemic warrant examination for possible bias in these models. We investigated the performance of three severity scoring systems (APACHE IVa, OASIS, SOFA) across ethnic groups in two large ICU databases in order to identify possible ethnicity-based bias.

**Method:** Data from the eICU Collaborative Research Database and the Medical Information Mart for Intensive Care were analysed for score performance in Asians, African Americans, Hispanics and Whites after appropriate exclusions. Discrimination and calibration were determined for all three scoring systems in all four groups.

**Findings:** While measurements of discrimination -area under the receiver operating characteristic curve (AUROC) -were significantly different among the groups, they did not display any discernible systematic patterns of bias. In contrast, measurements of calibration -standardised mortality ratio (SMR) -indicated persistent, and in some cases significant, patterns of difference between Hispanics and African Americans versus Asians and Whites. The differences between African Americans and Whites were consistently statistically significant. While calibrations were imperfect for all groups, the scores consistently demonstrated a pattern of over-predicting mortality for African Americans and Hispanics.

**Interpretation:** The systematic differences in calibration across ethnic groups suggest that illness severity scores reflect bias in their predictions of mortality.

**Funding:** LAC is funded by the National Institute of Health through NIBIB R01 EB017205. There was no specific funding for this study.

## Introduction

Severity scoring systems are employed in the intensive care unit (ICU) to perform severity adjustment for the purposes of benchmarking and research.^1^ It has generally been assumed that these systems are fair and objective in terms of their use across different ethnic groups. However, while it is known that such models may perform differently among disparate geographic populations or between different centres, ^2^ the assumption of scoring neutrality among ethnic groups within a given population has not been closely examined.

Disparities in ICU outcomes may result from pre-admission clinical factors, socioeconomic determinants, the quality of ICU care, and cultural practices.^3, 4^ Another possible source of disparity emanates from the use of biased algorithms.^5, 6, 7, 8^ The current COVID-19 pandemic raises two intersecting issues that demand a closer evaluation. First, relatively higher mortalities have been observed in particular ethnic populations, specifically African Americans.^9^ Second, severity scores have been proposed by professional societies and various policy groups to be incorporated into triage systems for potential scarce resource allocation.^10, 11^ It is therefore imperative to determine whether biased scoring systems could be adding to existent baseline disparities in healthcare.

The latest model of the Acute Physiology and Chronic Health Evaluation scoring system, APACHE IVa, was developed using data from 104 intensive care units in 45 USA based hospitals utilising 142 patient variables. The model employs the worst values in the first ‘APACHE day’ of the patient’s ICU stay to generate a risk score for hospital and ICU mortality and length of stay.^12^ The Oxford Acute Severity of Illness Score (OASIS) was developed from 81,087 admissions from 86 ICUs in the USA, utilising 10 variables collected in the first 24 hours of ICU stay.^13^ The Sequential Organ Failure Assessment (SOFA) score was developed based on expert opinion, incorporating organ function scores from six organ systems to characterize severity state in sepsis but has been repurposed to predict patient outcomes.^14^

In this retrospective observational study, we examined the performance of three severity scoring prediction models, namely APACHE IVa, OASIS, and SOFA in two large, publicly available ICU databases (eICU-Collaborative Research Database and Medical Information Mart for Intensive Care-III).

### RESEARCH IN CONTEXT

#### Evidence before this study

We searched PubMed on September 4, 2020 with no filter restrictions using the terms ‘intensive care unit severity scoring systems’ and ‘bias’ and ‘racial bias’ and found no results. These systems are used in critical care medicine for severity adjustment for research purposes and for benchmarking intensive care unit (ICU) performance. Ethnicity is generally documented in the process of hospital admission. However, none of the currently employed ICU severity scoring systems incorporate ethnicity or other relevant socioeconomic factors as a parameter in their analysis. We chose to examine three of these systems (APACHE IVa, OASIS, SOFA) for possible ethnically based bias. Out of these three, SOFA has come to be potentially employed (in guidelines) for initial ICU triage purposes and to determine the continuation of mechanical ventilation in situations of limited resources during a pandemic.

#### Added value of this study

We analysed the performance of three different clinical prediction models across four ethnicities in two large publicly available critical care databases involving 122, 919 and 43,823 admissions respectively. We found evidence that all three models over-predict mortality in all ethnic groups. While this general phenomenon of model drift is already known, we show that the over-prediction is more marked in African American and Hispanics, who are traditionally associated with poor socioeconomic status compared with Whites and Asians in the United States. This was consistent in both the databases for all the prediction models tested.

In view of the aforementioned use of one of the scoring systems (SOFA) in the current pandemic for purposes of triage of potentially limited resources and the disparate clinical outcomes of certain ethnic groups, we concluded that it is particularly important to ascertain whether severity scoring systems might contain previously undetected elements of bias which would make them inappropriate to utilize for clinical decision-making.

#### Implications of all the available evidence

Triaging of critical care resources is being discussed widely in the context of the current pandemic. In order to bring objectivity to the decision making, clinical prediction scores have been proposed to form part of the triage process. Sequential Organ Failure and Assessment (SOFA) is the most commonly proposed model in this context. We would maintain that we demonstrated sufficient evidence of bias in terms of the predicted versus observed mortalities (model calibration) that such use should be approached with extreme caution, and it may be most prudent to avoid applying these prediction models to critical care triage across populations involving patients from different socioeconomic and ethnic background.

## Methods

### The eICU Collaborative Research Database (eICU-CRD)

This eICU-CRD was derived from the eICU telehealth system.^15^ This system was designed to complement on-site ICU teams with remote support. The data include over 200,000 discharged patient episodes across 335 ICUs at 208 hospitals during the period of 2014-2015. Patient demographics including age, sex, ethnicity, vital signs, diagnoses, laboratory measurements, clinical history, problem lists, APACHE IVa scores and treatments are available in the database.

### MIMIC-III database

MIMIC-III (Medical Information Mart for Intensive Care III) is a publicly available database consisting of over 60,000 ICU admissions to the Beth Israel Deaconess Medical Centre (BIDMC) between 2001 and 2012. ^16^ MIMIC-III incorporates OASIS as a mortality prediction model.

Admission SOFA scores were computed in both databases. Mortality in all ethnic groups were calculated at multiple SOFA cut-offs, with SOFA score categories of 0-7, 8-11 and >11. The categories are based on what have been proposed for COVID-19 ventilator allocation. ^10^

The Federal guidance in the USA classifies race into five categories, and ethnicities into two categories.^17^ For this paper, we defined “ethnicity” based on the entries made in the demographic sections of the respective databases. The ethnicities included in the analyses were African American, Asian, Hispanic and White. Native Americans were excluded due to the much smaller sample size compared to the other ethnicities (n=946 (0.7%) in eICU and n=57 (0.11%) in MIMIC). Patient episodes with a nonspecific or unknown ethnicity category were excluded.

Patients with missing survival data or erroneous/missing prediction scores, missing ethnicity data and those <16 years or >90 years of age were excluded from the analyses.

Ethnicity information was available in both the databases. It is typically entered by an administrator who asks the patient or the family member which ethnic group they identify with, or obtained from previously available records.

### Tests of discrimination

Discrimination was determined by the Area Under Receiver Operating Characteristic (AUROC) curve for different ethnic groups. Hospital mortality was the outcome of interest. SOFA score was analysed in both databases, while APACHE IVa and OASIS were used as predictors in the eICU-CRD and MIMIC-III databases respectively.

### Tests of calibration

Calibration was evaluated using standardised mortality ratio (SMR) for APACHE IVa and OASIS. Since predicted mortality for a given SOFA score for an individual patient cannot be calculated, SMR could not be specifically calculated for SOFA. Instead, observed mortality for each ethnic group was compared to the mortality rate in the overall population in that SOFA score category in order to provide an evaluation of comparative outcomes among ethnic groups. To further characterise model performance in the context of sicker patient populations, an additional calibration analysis was performed across risk grades of 0-5%, >5-10%, >10-20%, >20-50% and >50%, based on APACHE IVa and OASIS in eICU and MIMIC-III patients respectively. (*Supplementary appendix*)

The statistical analyses were performed in R v4.0.0. The packages used included: rsq (partial R2) – v2.0; ems (SMR) -v1.3.2; dplyr (data handling and summarising) – 1.0.0 and pRoc. Stata version 14 was used for comparison of AUROC between groups using the Roccomp function.

### Ethical approval

Research using the eICU-CRD is exempt from institutional review board (IRB) approval due to the retrospective design, lack of direct patient intervention, and the security schema, for which the re-identification risk was certified as meeting safe harbour standards by an independent privacy expert (Privacert, Cambridge, MA) (Health Insurance Portability and Accountability Act Certification no. 1031219-2). The data in MIMIC-III has been previously de-identified, and the IRBs of the Massachusetts Institute of Technology (No. 0403000206) and BIDMC (2001-P-001699/14) both approved the use of the database for research.

## Results

The distribution and characteristics of patients in different ethnic groups are shown in Table 1. As shown in Figure 1, the following patients were excluded: missing/unknown ethnicity, ethnicity outside the 4 being examined, those outside the 16-89 age range and those without a valid model predicted mortality (required for SMR calculation). The total numbers of ICU admissions included in the final analysis were 122,919 (83.8% of all episodes) and 43,823 (71.2% of all episodes) in the eICU-CRD and MIMIC-III respectively.

**Table 1:**
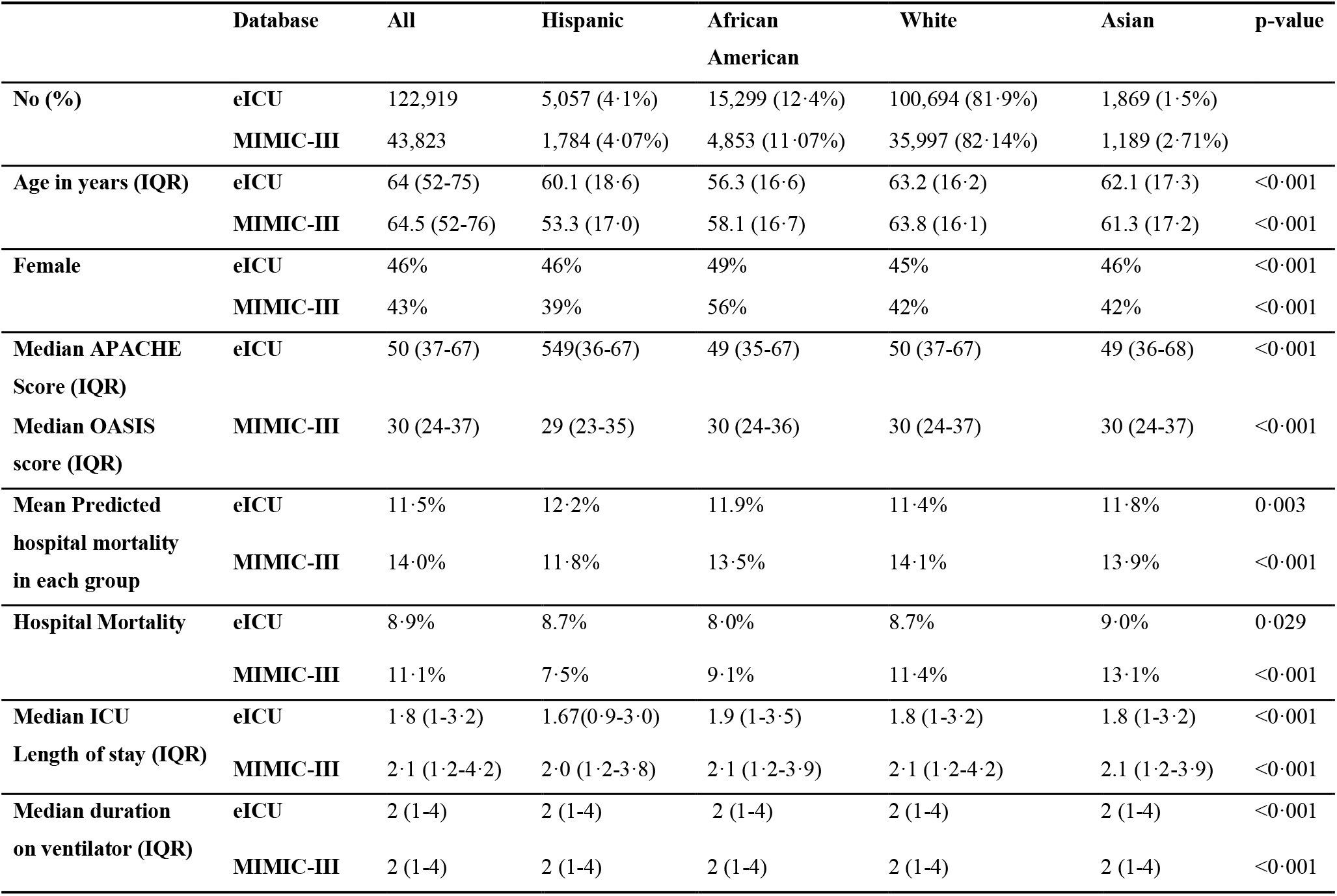
Patient characteristics across the ethnic groups

**Figure 1:**
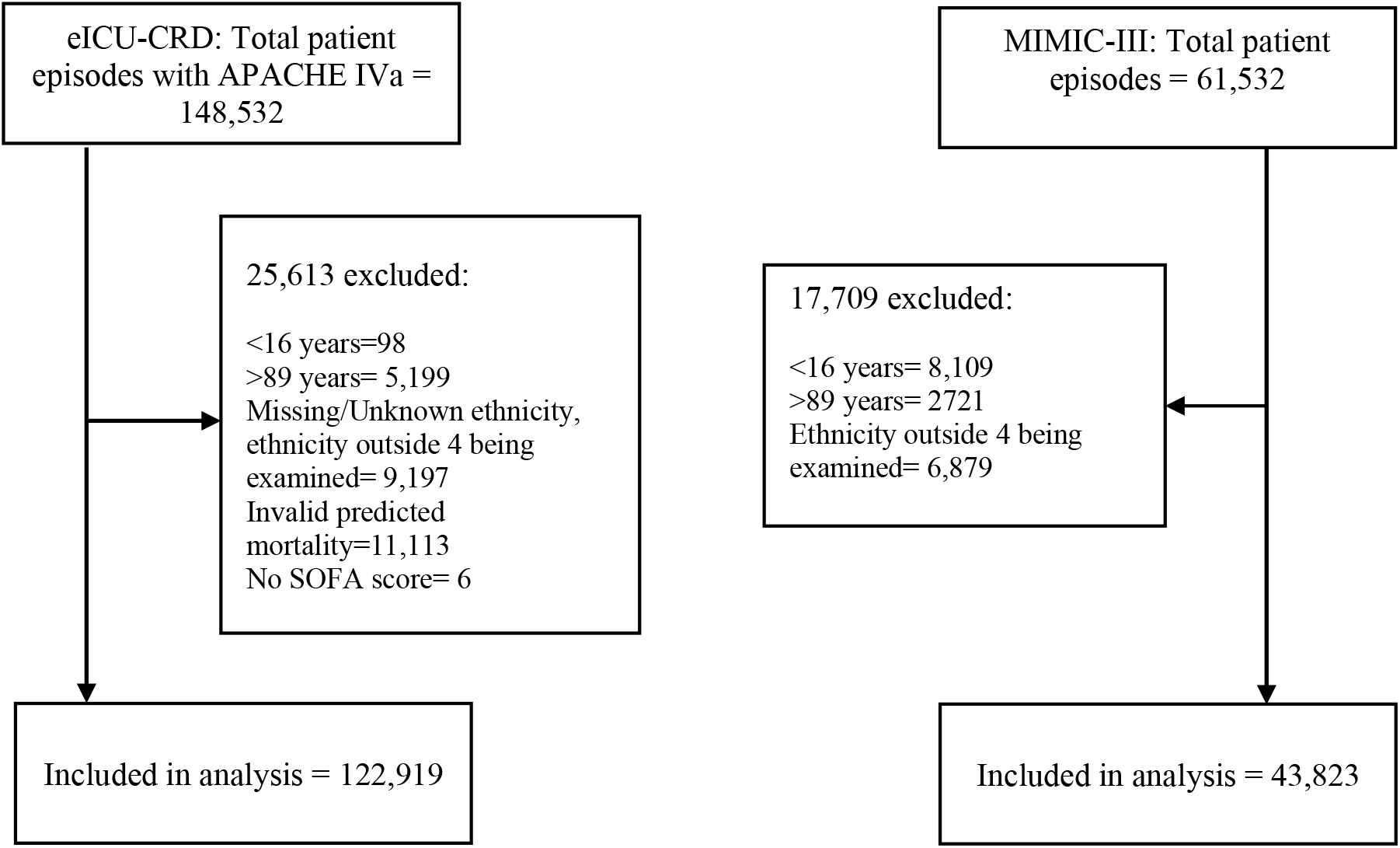
**E**xcluded patients in both databases; the exclusions have been made in the sequence specified in the diagram.

African Americans and Hispanics were younger than patients of other ethnicities. Mean prediction scores were similar across the groups. Predicted hospital mortalities across ethnicities were in the 11%-12% range in the eICU-CRD and 11%-14% in MIMIC-III, while observed mortalities were 8%-9% in the eICU-CRD and 7%-13% in MIMIC-III, indicating that both models over-estimated hospital mortality.

### Discrimination

Tests for discrimination showed that the APACHE IVa model performed well across all ethnic groups in the eICU-CRD, with the AUROC, for the Hispanic, African American, Asian and White groups being 0·89, 0·87, 0·86 and 0·86, respectively (See Table 2 and Figure 2). Across group differences in the AUROC were statistically significant (p=0·016). The AUROCs in MIMIC-III in the Hispanic, African American, White and Asian groups were 0·76, 0·75, 0·76 and 0·77, respectively, displaying insignificant across group differences (p=0·85).

**Table 2:** AUROC in different ethnicities in both the databases (with 95% CIs)

| Scoring system (Database) | Hispanic | African American | White | Asian | p-value |
| --- | --- | --- | --- | --- | --- |
| APACHE IVa (eICU) | 0.88<br>(0.8744, 0.903) | 0.87<br>(0.857, 0.878) | 0.86<br>(0.860, 0.868) | 0.86<br>(0.831, 0.889) | 0.016 |
| OASIS (MIMIC-III) | 0.76<br>(0.72,0.81) | 0.75<br>(0.73,0.78) | 0.76<br>(0.75,0.77) | 0.77<br>(0.73, 0.81) | 0.85 |

**Figure 2:**
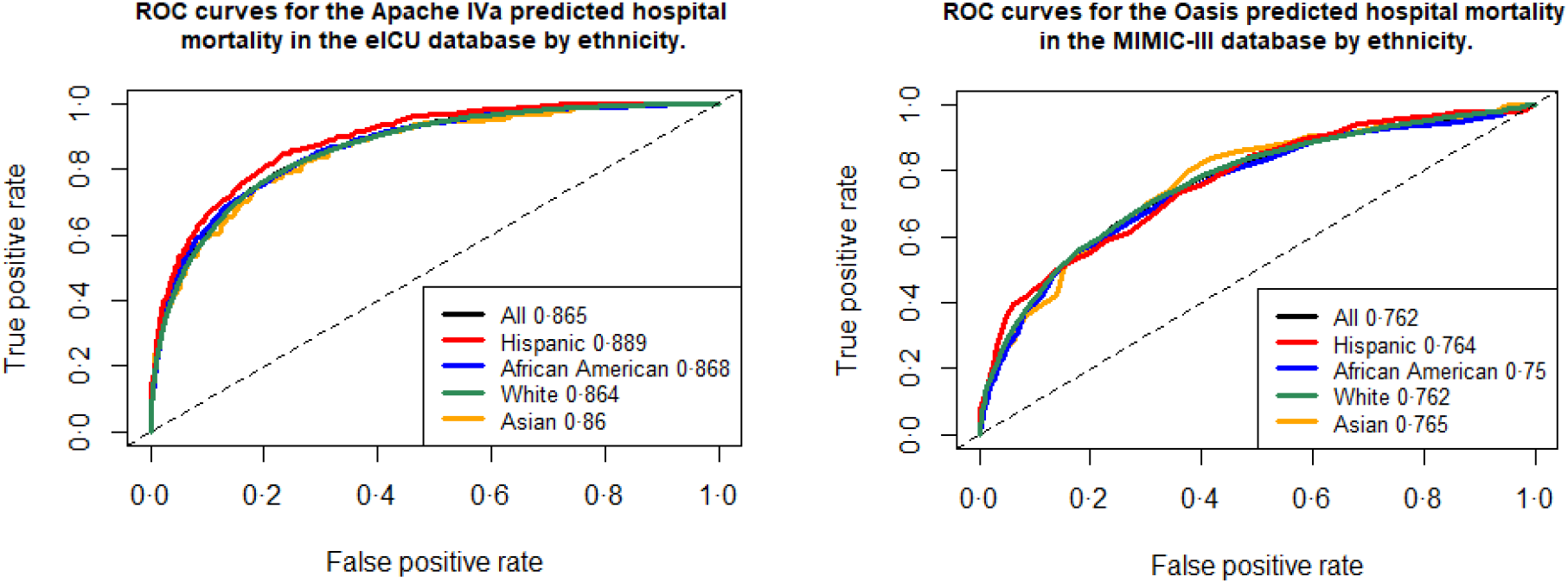
**Left panel:** AUROCs in different ethnic group in eICU for APACHE IVa. **Right panel**: AUROC in different ethnic group in MIMIC-III for OASIS.

### Calibration

The observed/predicted death rates in the eICU-CRD were 442/608, 1219/1813, 8732/11456, and 169/220 in the Hispanic, African American, White, and Asian groups, respectively. Therefore, across all groups, APACHE IVa predicted more deaths than were actually observed, with SMRs of 0·73, 0·67, 0·76, and 0·77 in the same group order (Table 3). This over-prediction of mortality was also observed in MIMIC-III, with the SMR values of Hispanics, African Americans, Whites and Asians being 0·64, 0·67, 0·81 and 0·95 respectively. As can be seen in Table 3 and Figure 4, the APACHE IVa model was least accurate for predicting hospital mortality in African Americans (SMR 0·67) and most accurate in Asians (SMR 0.77). OASIS was least accurate in Hispanics and African Americans (SMRs of 0·64 and 0·67 respectively), and most accurate in Asians (SMR 0·95). Forest plots of the SMR data are displayed in Figure 4. While all the differences do not quite reach statistical significance, there do appear to be two distinct groupings consisting of Hispanic/African American patients, and Asian/White patients, with the former displaying significantly worse calibration than the latter. The difference in calibration between the African American/Hispanic groups and the White/Asian groups is significant in MIMIC-III.

**Table 3:** Predicted and observed mortality in different ethnic groups along (SMR = actual/predicted mortality ratio in each patient group); SMRs in eICU are for APACHE IVa and SMRs for MIMIC-III are for OASIS scores respectively.

| Ethnicity | Dataset | Total number | Mean predicted mortality | Predicted deaths | Actual deaths | SMR |
| --- | --- | --- | --- | --- | --- | --- |
| Hispanic | eICU | 5057 | 0.12 | 608 | 442 | 0.73 |
|  | MIMIC-III | 1784 | 0.12 | 210 | 134 | 0.64 |
| African American | eICU | 15299 | 0.12 | 1813 | 1219 | 0.67 |
|  | MIMIC-III | 4853 | 0.14 | 657 | 443 | 0.67 |
| White | eICU | 100694 | 0.11 | 11456 | 8732 | 0.76 |
|  | MIMIC-III | 35997 | 0.14 | 5081 | 4114 | 0.81 |
| Asian | eICU | 1869 | 0.12 | 220 | 169 | 0.77 |
|  | MIMIC-III | 1189 | 0.14 | 165 | 156 | 0.95 |
| p-value (across group difference) | eICU |  |  |  |  | <0.0001 |
|  | MIMIC |  |  |  |  | <0.0001 |

**Figure 3:**
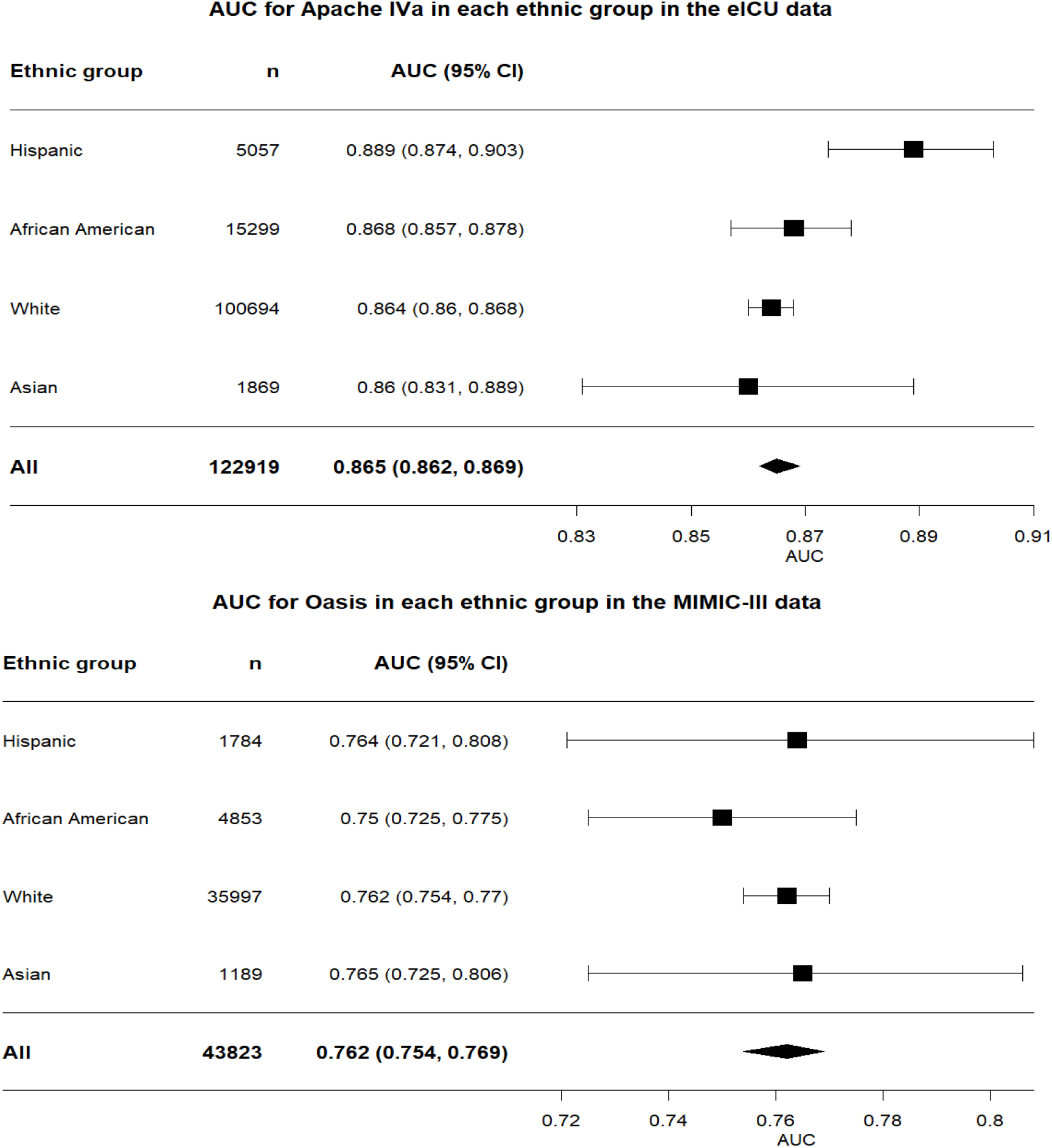
**Top panel:** Forest plot for AUROCs from the eICU-CRD. There is a clear separation between the White and Hispanic groups. All other group pairs have overlapping 95% confidence intervals (CIs). **Bottom panel:** Forest plot for AUROCs from the MIMIC-III. The absolute AUROCs are similar, with overlapping 95% CIs for all groups.

**Figure 4:**
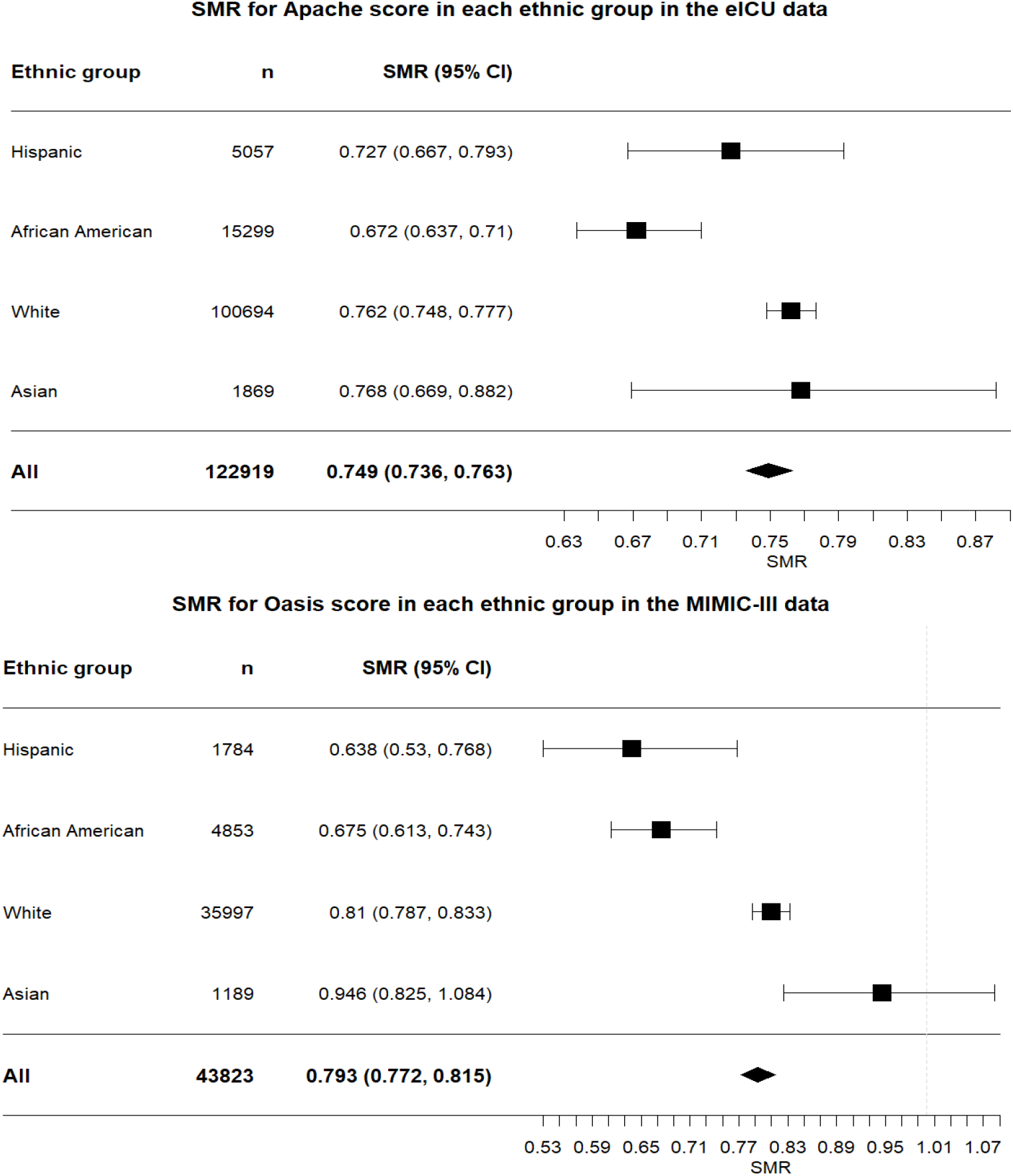
**Top panel:** Forest plot for SMRs for different ethnic groups from the eICU for mortality predicted by APACHE IVa. There is a clear separation between White and African American groups. However, all other groups have overlapping 95% confidence intervals, with a lower SMR point estimate for Hispanics, compared to Whites and Asians. **Bottom panel**: Forest plot for SMRs for different ethnic groups from the MIMIC-III for predicted mortality determined by OASIS. There is clear separation of CIs between the African American/Hispanic and White/Asian groups. Hispanic and African American SMRs again are lower than Asian and White.

In all the ethnic groups, SMR was higher and generally closer to 1 with increasing predicted risk categories, signifying that APACHE IVa and OASIS are better calibrated across all ethnicities in sicker patient populations. *(See the supplementary appendix)*.

### Performance of SOFA in the eICU-CRD and the MIMIC-III

Discrimination in both the databases was comparable across ethnicities, with the exception of Asians in the eICU-CRD where the AUROC was considerably lower (See Figure 5). For the other three ethnic groups in this database, AUROCs ranged between 0·767 and 0.787. In the MIMIC-III, AUROCs ranged between 0·73 and 0·757. As we noted in the methods, usual SMRs could not be calculated to determine calibrations for SOFA. However, using the approach we described, we observed the same phenomenon of a lower observed mortality for a given risk score category in African Americans (and less so for Hispanics), compared with Whites and Asians across many of the score categories (Table 4 and Table 5). SOFA mortalities were also notably different between the databases for the same scoring category within a given ethnic group.

**Table 4:** Proportion(%) of mortality in different admission SOFA score ranges across ethnic groups in the eICU and MIMIC-III.

| Initial SOFA score | Database | All patients | Hispanic | African American | White | Asian | p |
| --- | --- | --- | --- | --- | --- | --- | --- |
| 0-7 | MIMIC-III (n=38,011) | 7.6 | 4.7 | 5.6 | 7.9 | 9.6 | <0.0001 |
|  | eICU (n=110,671) | 6.0 | 5.79 | 5.17 | 6.12 | 6.73 | <0.0001 |
| 8-11 | MIMIC-III (n=4,609) | 27.7 | 18.2 | 24.4 | 28.5 | 29.6 | 0.004 |
|  | eICU (n=10207) | 27.63 | 28.64 | 26.38 | 27.76 | 28.89 | 0.65 |
| >11 | MIMIC-III (n=1,203) | 57.2 | 61.2 | 56.8 | 57.2 | 54.1 | 0.92 |
|  | eICU (n=2041) | 54.24 | 58.7 | 49.5 | 54.8 | 57.69 | 0.20 |

**Table 5:** Ratio of observed mortality to overall mortality by admission SOFA category and ethnic group in both databases.

| SOFA score | Database | Hispanic | African American | White | Asian |
| --- | --- | --- | --- | --- | --- |
| 0-7 | MIMIC | 0.62 | 0.74 | 1.04 | 1.6 |
|  | eICU | 0.96 | 0.86 | 1.02 | 1.12 |
| 8-11 | MIMIC | 0.66 | 0.88 | 1.03 | 1.07 |
|  | eICU | 1.04 | 0.95 | 1.00 | 1.05 |
| >11 | MIMIC | 1.07 | 0.99 | 1 | 0.95 |
|  | eICU | 1.08 | 0.91 | 1.01 | 1.06 |

**Figure 5:**
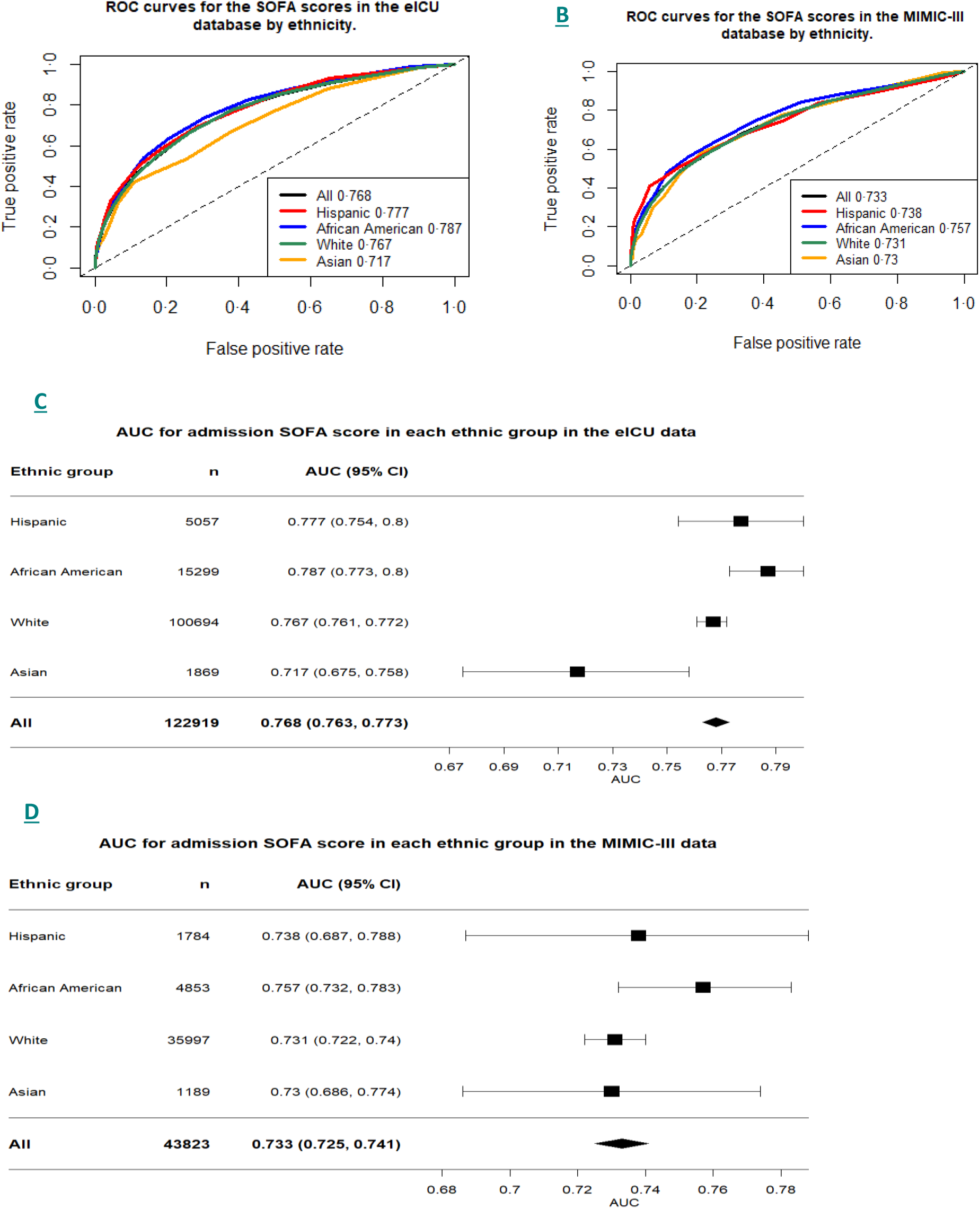
**Panel A:** AUROC plots for all ethnicities in the eICU-CRD for SOFA score performance in hospital mortality prediction. **Panel B:** AUROC plots for all ethnicities in MIMIC-III for SOFA score performance in hospital mortality prediction. **Panel C**: Forest plot for AUROCs in different ethnicities in the eICU-CRD for performance of SOFA score with 95% confidence intervals, showing Asians to be an outlier. **Panel D:** Forest plot for AUROCs in different ethnicities in MIMIC-III for performance of SOFA scores with 95% confidence intervals. This shows overlapping of 95% confidence intervals of AUROCs across all groups.

## Discussion

In this first comparative study of ICU mortality prediction model performance in different ethnicities, we show that while there was a statistically significant difference across the AUROCs, there was no systematic pattern to the difference in the discriminative performances of APACHE IVa, SOFA, and OASIS. However, with regard to calibration, OASIS, APACHE IVa, and SOFA over-predicted mortality in all ethnic groups. This poor calibration was particularly notable in the African American and Hispanic groups. There was a statistically significant difference between the SMRs of Whites and African Americans for both APACHE IVa and OASIS, and a statistically significant difference between Whites and Hispanics for OASIS. Asians were statistically different from African Americans and Hispanics in OASIS only. (Figure 4)

Although not designed for mortality prediction, SOFA performed reasonably well in terms of discrimination with the exception of the somewhat aberrant AUROC in the Asian group in eICU-CRD. The relative mortality risks in Hispanics and African Americans were notably different (lower) in the two databases for low to moderately high SOFA scores. This must be taken into consideration when SOFA is employed for prognostication and triage decisions in the ICU.

Importantly, while it is reassuring that all scores were better calibrated in the sicker population, it is of concern that in mild to moderate risk categories, including mid-range SOFA scores, calibration is poor in the ethnic groups who are usually associated with poor socioeconomic backgrounds. While calibrations were less disparate at the highest score levels (indicating very poor prognoses) of >11, the mortality ratio for African Americans was still >10% lower than that of Whites and Asians in the ICU database at this level.

These findings have potential repercussions for some of the guidelines on the appropriation of limited ICU resources during the pandemic. It has been proposed, for example, that for a persistent SOFA score of 8-11 after 48 or 120 hours, treatment continuation should be evaluated. ^10, 11^ If SOFA does indeed over-predict mortality in that score range, then this form of decision making could be misguided. The same guidelines from New York and Michigan have used a level of 12 as a potential cut-off for admission or continued ICU care. The critical question raised by this study is why the African American and Hispanic groups demonstrated such inaccurately high mortality predictions. In this context, the most concerning potential scenario would be withholding of treatment or withdrawal of care on the basis of a perhaps falsely high predicted mortality.

Precise calibration is particularly important if these systems are to be used for care decisions in individual patients. Triage decisions related to patient admission, management (including discontinuation of treatment), and discharge from the ICU are potentially subjective and vulnerable to bias. Scoring systems may be applied to these decisions in order to, in theory, introduce a greater level of objectivity and fairness when resources are critically limited. However, if the systems themselves are biased, then their use for these purposes will systemically imprint and effectively endorse existing inequities. Another important point is the utilisation of prediction models based on a single time point, as this may not always capture an individual patient’s potential to respond to a proposed treatment. However, in real-world decision making, especially in a resource constraint scenario, all the clinician or a triage official has is a snapshot type of risk prediction tool.

While a temporal drift in model performance may explain low SMRs in all the ethnic groups, it is not clear why these scoring systems produce ethnically consistent patterns of poor calibration. Based on recent papers, it is unlikely that African American and Hispanic patients received relatively better care. ^18, 19^ It is also doubtful that an identical physiological phenotype represents a different disease trajectory in those groups. An implicit assumption of scoring systems is that patients have the same baseline states and that the scores represent the same degree of deviation from that baseline state. However, African Americans and Hispanics admitted to an ICU with the same severity scores as Whites and Asians, may actually be exhibiting a smaller change from their baseline status. For example, a population with a higher prevalence of chronic organ failure (e.g. baseline elevations in serum creatinine or bilirubin) could demonstrate SOFA scores that do not accurately portray their acute physiological status. Deliberato et al. demonstrated that patients with obesity, which may well be higher in the African American and Hispanic populations,^20^ may be similarly misclassified with regard to illness severity based on absolute physiologic measurements on ICU admission given a more abnormal baseline state compared to patients with a lower body mass index.^21^ In the same vein, it has been shown that the chronic disease burden may contribute more towards mortality in critically ill.^22^ Given that the Hispanics and African-Americans were younger than the other two ethnic groups in both the databases, it is possible that they had a low chronic disease burden, resulting in a lower contribution of chronic disease towards mortality risk for the similar acute physiological profile.

In a perfect world without bias and health disparities, only patient and disease factors would determine case-mix and clinical outcomes in the ICU. However, studies have repeatedly demonstrated that this is far from the case.^18, 19^ Our detection of inadvertent but undeniable bias in severity scores would seem to indicate that it is time to develop scoring systems that are more precise than the current ‘one size fits all’ systems. This will admittedly pose a challenge, but one that is achievable as more data accumulate for varying patient cohorts and contexts. In response to this need, there is a movement across the critical care community to make mortality risk prediction models more dynamic and useful in real time, often based on data collected from electronic health records.^23, 24, 22, 25, 26, 27^ Notably, around 70% of the patients were White in the training and validation datasets for APACHE and OASIS models. More diverse ethnic representation of patients during model development will help reduce potential bias. Attention must also be paid to relevant sociodemographic factors while developing the models. Especially, with the potential resource limitations arising in the COVID-19 pandemic, the wide use of biased risk prediction models is undoubtedly problematic.^28^ Access to care, including life-saving treatments, is the strongest predictor for, and a potential root cause of, poor health outcomes.^29^ Evidence also exists that health outcomes differ significantly within an ethnic group depending on income and education.^30, 31^

To add to the complexity surrounding this issue, there persists a debate whether race is a social or a biological concept.^32^ In fact, there are greater genetic differences between individuals of the same ethnic group than there are differences across ethnic groups. Furthermore, because socioeconomic factors may be distributed disproportionately, it has been recommended that both ethnicity and socioeconomic parameters are included in health reporting.^30, 33^ A mere race adjustment may further the disparity in care.^34^

In addition to their use for triage purposes, these scoring systems are used for severity adjustment in research and for benchmarking performance. Our findings will need to be taken under consideration for these purposes, as well. For example, an ICU with a largely African American population would appear to be performing better than a unit of largely White patients on the basis of model mortality overpredictions for the African Americans. For research, populations thought to be of equal severity, may not be quite so. These are important considerations that will need to be addressed, but not of the urgency of the potential bias of systems employed for triage purposes. Another important point is that given that MIMIC-III and eICU capture a wide variety of ICUs in the U.S., these data should be potentially generalisable to most western settings where triaging of critical care resources on the basis of risk prediction tools have been discussed. However, a local assessment of model performances in different ethnic group in different settings is needed.

There are a number of limitations of our study. First, there was elimination of patients with missing ethnic data from the analysis. Missing data is unfortunately an integral part of real-world clinical data analysis, and although extremely unlikely to be due to systematic bias, it is not possible to ascertain what resulted in the absence of the ethnic data in those patients. Second, the ascertainment of ethnicity was done at individual hospitals and was largely based on self-reporting. Third, the attribution of certain score components (e.g. Glasgow Coma Scale) could be somewhat subjective. However, this issue is an inherent nature of ICU risk scoring and would be a factor in any study of similar nature. Fourth, the ethnic group category for Asians is very heterogeneous including Indian-Asians, Filipino-Asians, Chinese-Asians and others. There may be significant differences to the performance of the scoring systems in these sub-groups that would be lost after aggregation.

In conclusion, we found that the APACHE IVa, SOFA, and OASIS predictive models performed discrimination in a manner that was technically but not systematically different between ethnic groups. However, all of these prediction models significantly and systematically overestimate mortality across all ethnic groups. Importantly, this poor level of calibration was most notable in Hispanic and African American patients and was found in all three scoring systems. In a world with health disparities whose healthcare providers’ triage decisions may be tainted with bias, current severity scoring prediction models may not be able to correctly and fairly characterize patient severity and risk. Incorporating precise socioeconomic and geographic parameters along with a set of specific biomarkers for a given disease into future prediction models may potentially make such models less biased and therefore, more robust. Extreme care must be taken in the application of current scoring systems for triage decisions in individual patients, if they are, in their present states, to be used at all for these purposes.

## Data Availability

The underlying data used was the MIMIC-III and eICU data hosted by Physionet.org. It is available for bona-fide research projects on completion of appropriate training of investigators. The code used in the analysis is available on GitHub.

https://github.com/cjmartin0/ITUscoringAnalysis

## Author contribution

RS: Conceived the study, literature search, study design, drafting and reviewing the manuscript; CM: Statistical analysis, reviewing the manuscript; HM: Critical review of the manuscript; JWG: Critical review of the manuscript; DJS: Study design, Drafting and reviewing of manuscript; LAC: Conceived the study, study design, writing and reviewing the manuscript. RS and CM have accessed and verified the data reported in the manuscript.

## Data sharing

The MIMIC-III and eICU Collaborative Research databases are publicly available through PhysioNet (www.physionet.org). Materialised views for the SOFA calculation are available in the respective code repositories. The code used for analysis can be found on GitHub here: https://github.com/cjmartin0/ITUscoringAnalysis (github.com).

## Supplementary appendix

Summary of the three prediction scores in the study:

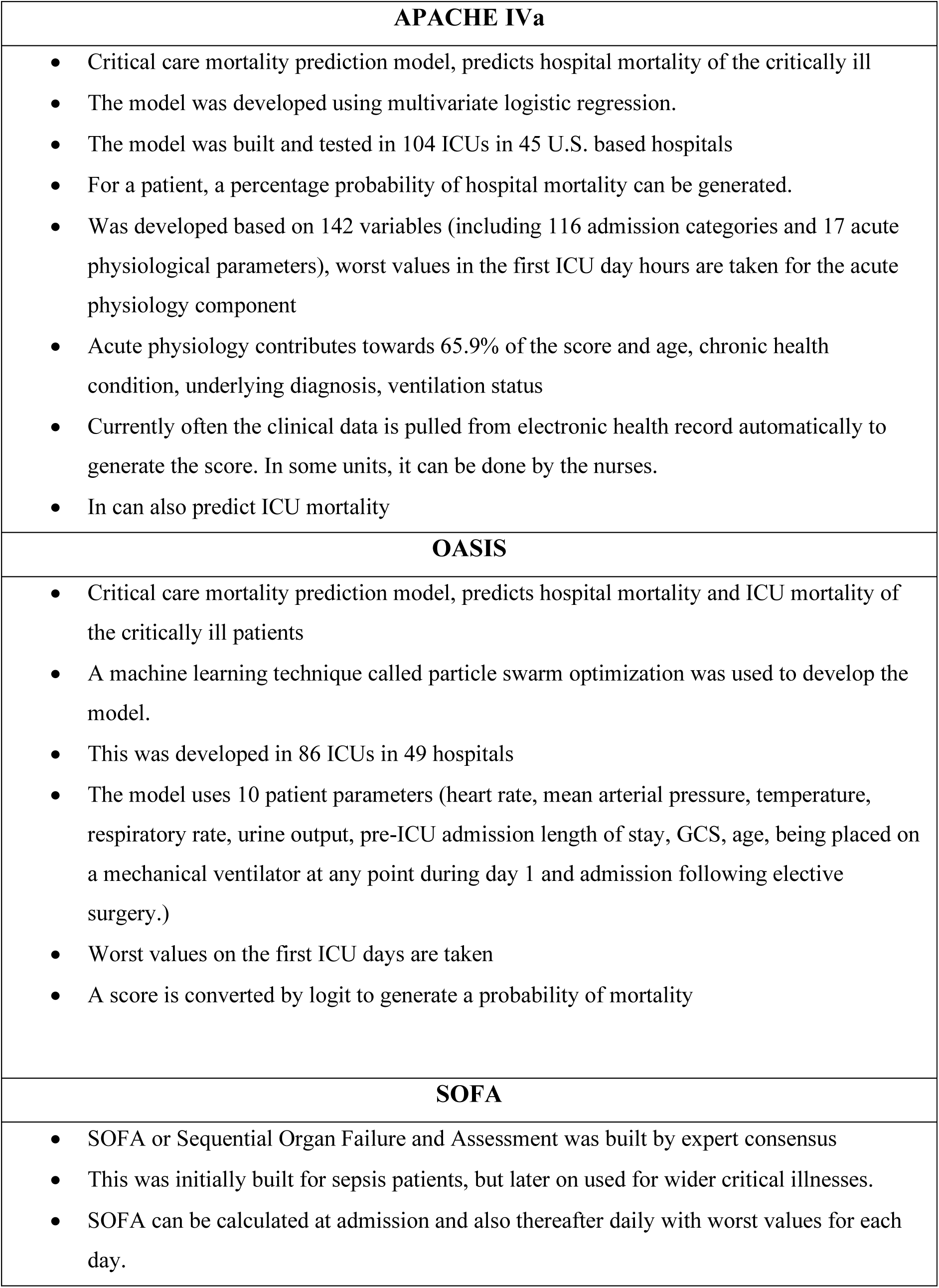
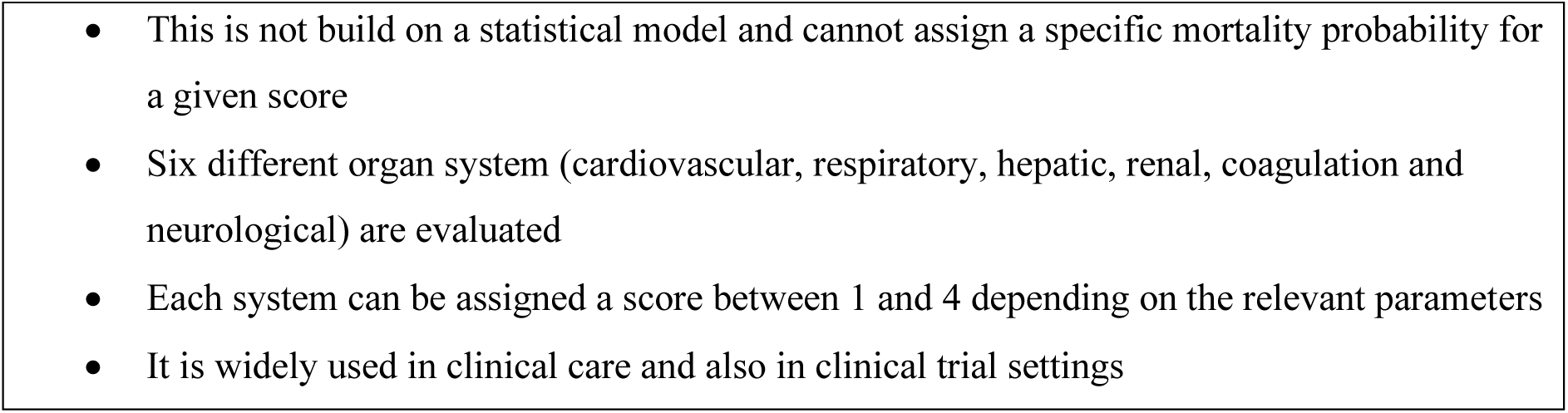

### Additional discussion on risk categories of APACHE IVa and OASIS

Table S1 shows the trend of increasing SMR with increasing predicted mortality risk. This was the same in all ethnic groups, as shown in Figures S1 and S2. In the lower risk categories, SMR was markedly low in certain groups. For example, SMR was 0·47 in Hispanics in MIMIC-III within the 10-20% risk category. This pattern was the same in other risk strata in the eICU data as well (e.g. African Americans and Hispanics had SMRs of 0·49 and 0·31 respectively in the eICU data’s 0-5% risk category). An exception to this trend of improving SMR with increasing predicted risk was African Americans in MIMIC-III, where no such improvement was seen and the patients within this group had persistently low SMRs (<0·7) in all risk categories.

**Supplementary table S1:** SMR across risk categories in different ethnic groups

| **Risk categories** | **Dataset** | **Hispanic** | **African American** | **White** | **Asian** |
| --- | --- | --- | --- | --- | --- |
| **0-5%** | eICU | 0.31 | 0.49 | 0.51 | 0.54 |
|  | MIMIC | 0.55 | 0.72 | 0.95 | 0.65 |
| **5-10%** | eICU | 0.66 | 0.65 | 0.69 | 0.76 |
|  | MIMIC | 0.63 | 0.72 | 0.75 | 0.67 |
| **10-20%** | eICU | 0.72 | 0.61 | 0.77 | 0.89 |
|  | MIMIC | 0.47 | 0.54 | 0.74 | 0.92 |
| **20-50%** | eICU | 0.71 | 0.67 | 0.79 | 0.65 |
|  | MIMIC | 0.64 | 0.67 | 0.8 | 0.85 |
| **50-100%** | eICU | 0.90 | 0.77 | 0.85 | 0.90 |
|  | MIMIC | 0.92 | 0.68 | 0.86 | 0.96 |

**Supplementary figure 1:**
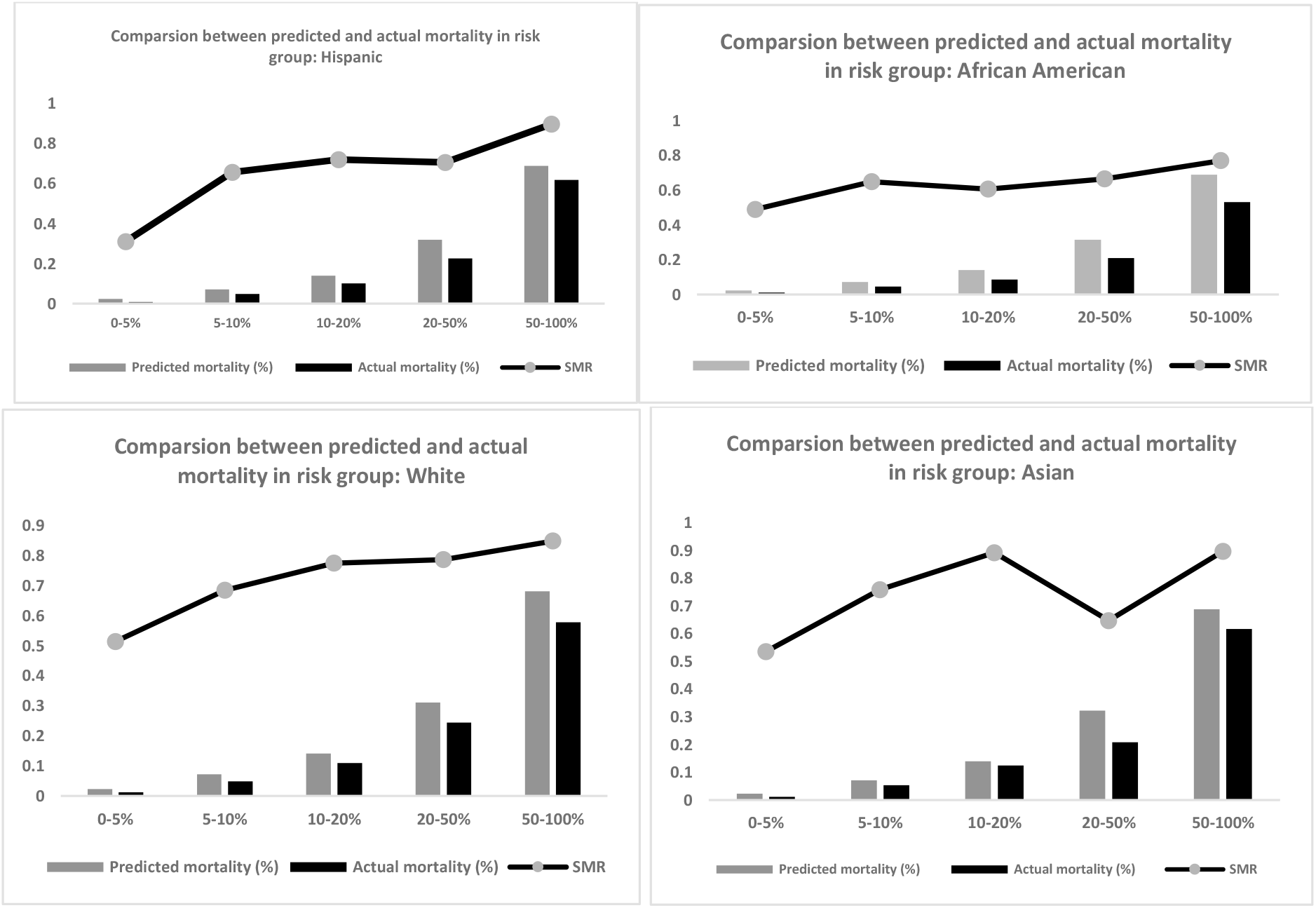
SMR trend in different risk categories in all ethnic groups in eICU database

**Supplementary figure 2:**
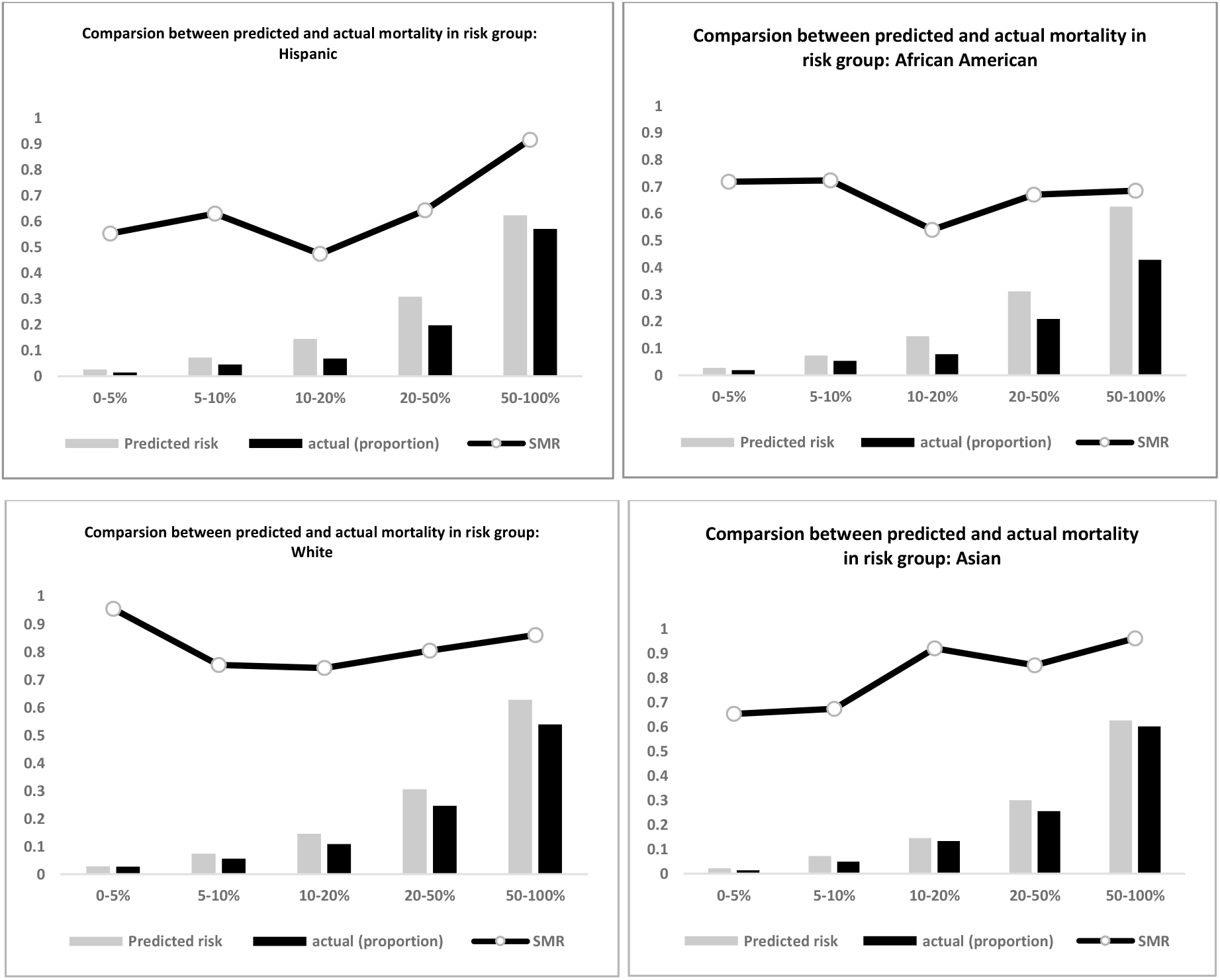
Trend in different risk categories in all ethnic groups in MIMIC-III database

**Supplementary figure 3:**
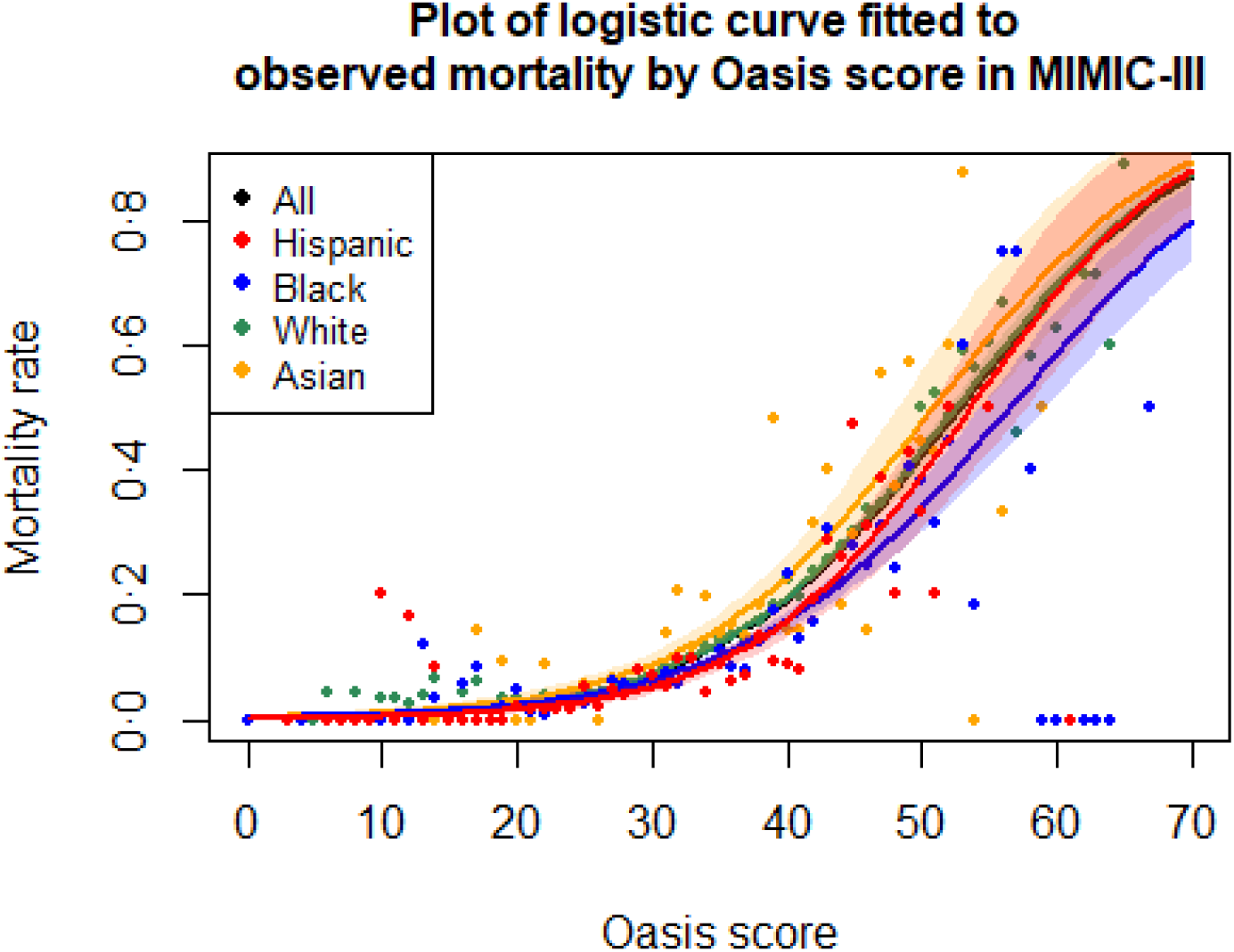

**Supplementary figure 4:**
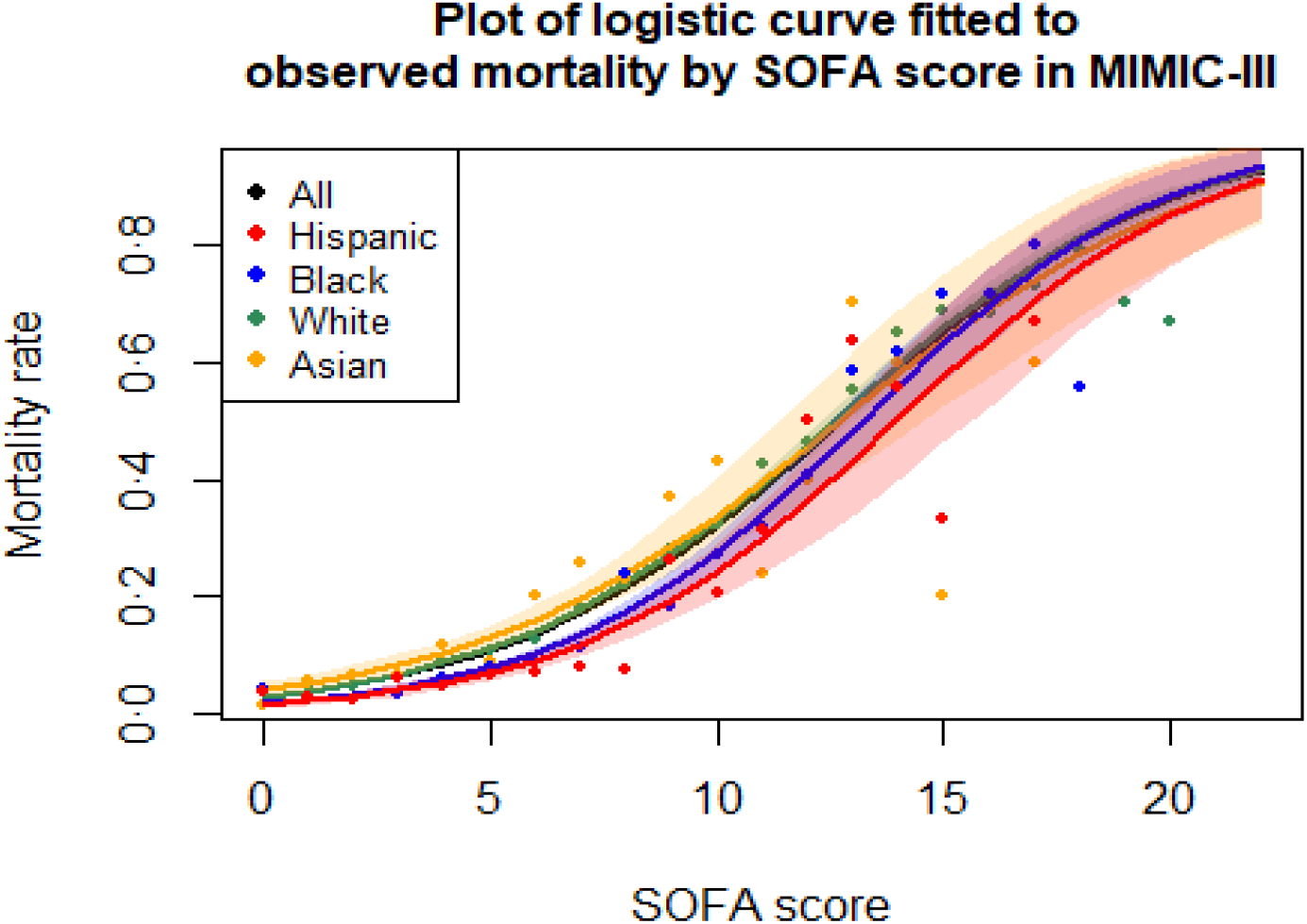

**Supplementary figure 5:**
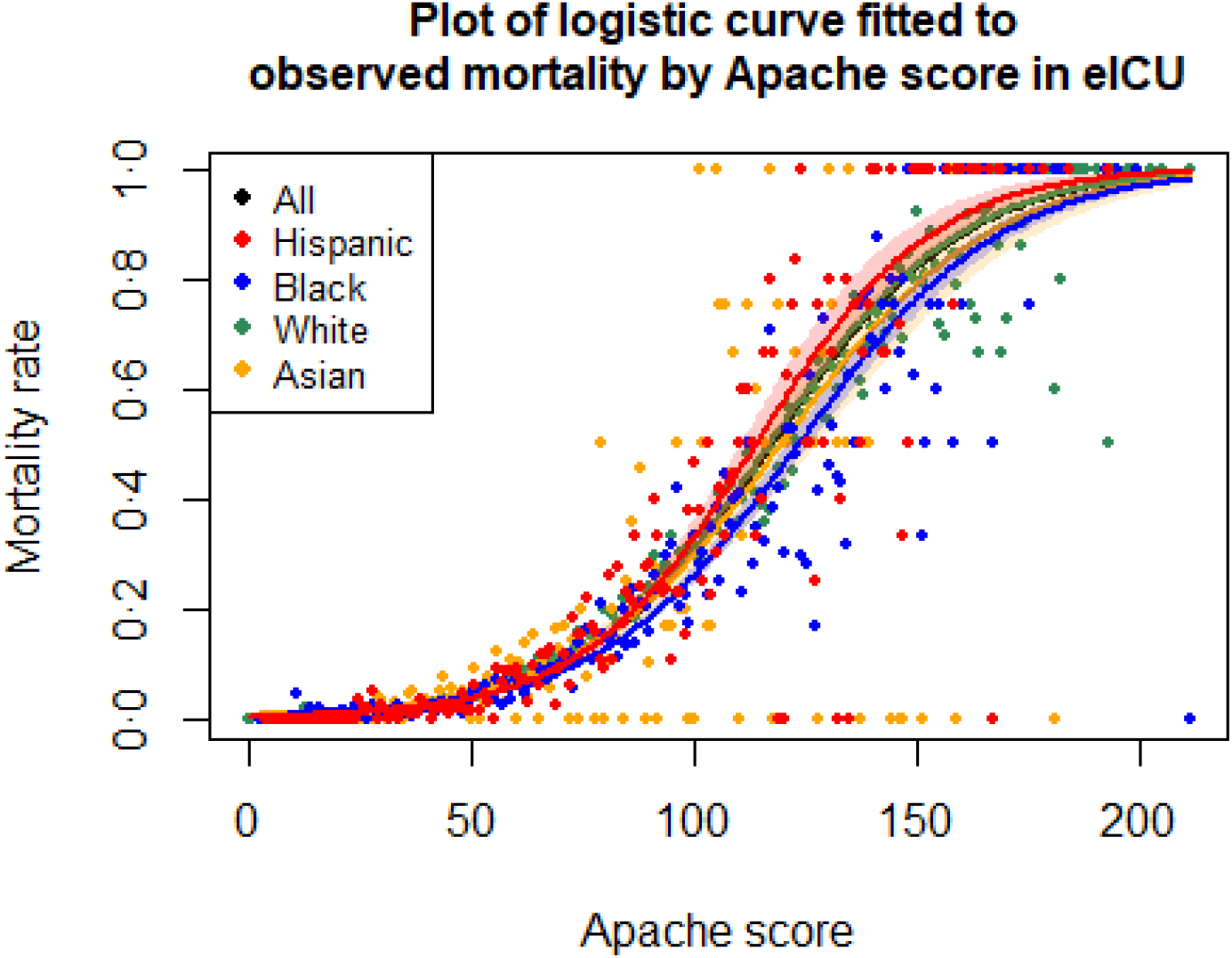

**Supplementary figure 6:**
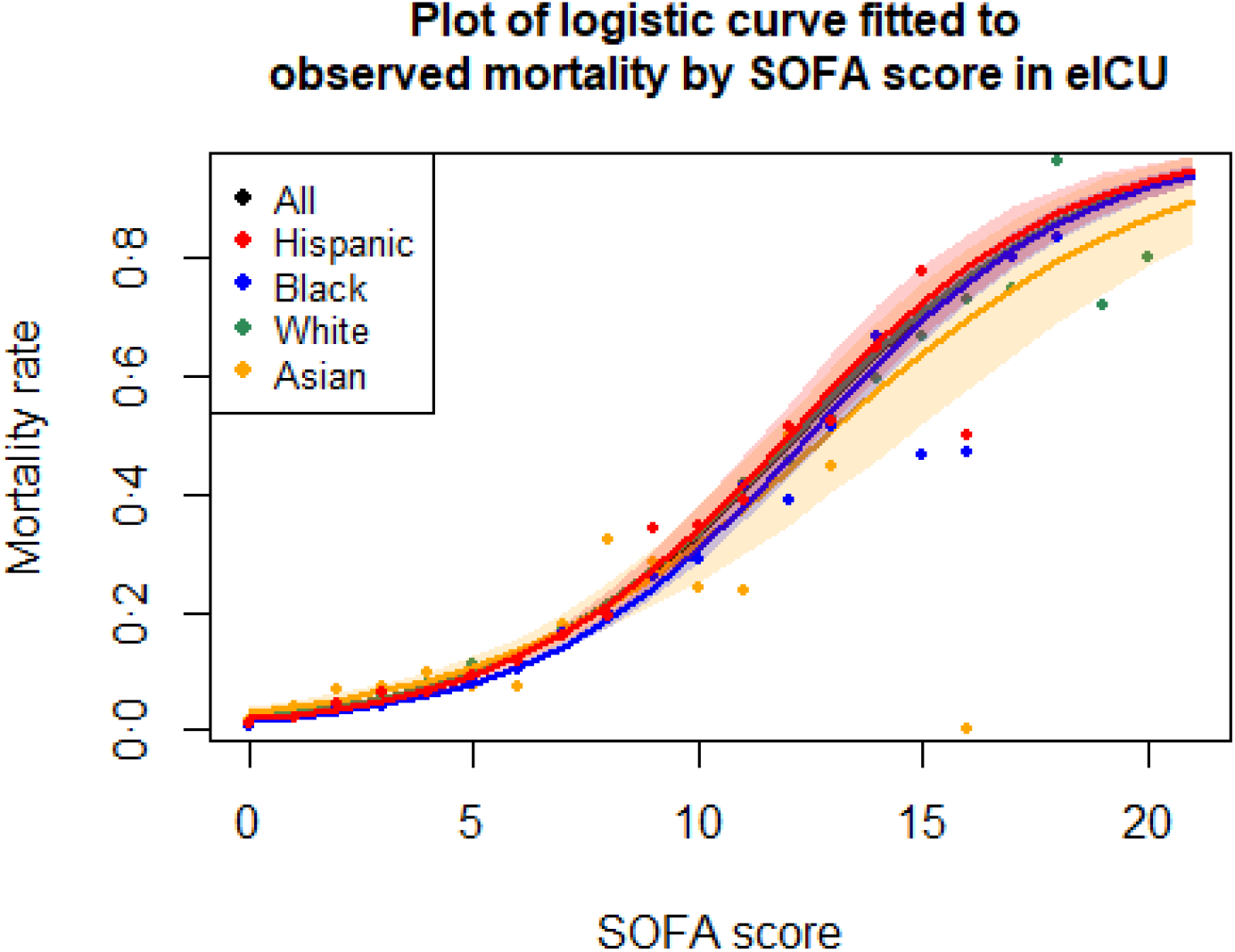

**Supplementary figure 7:**
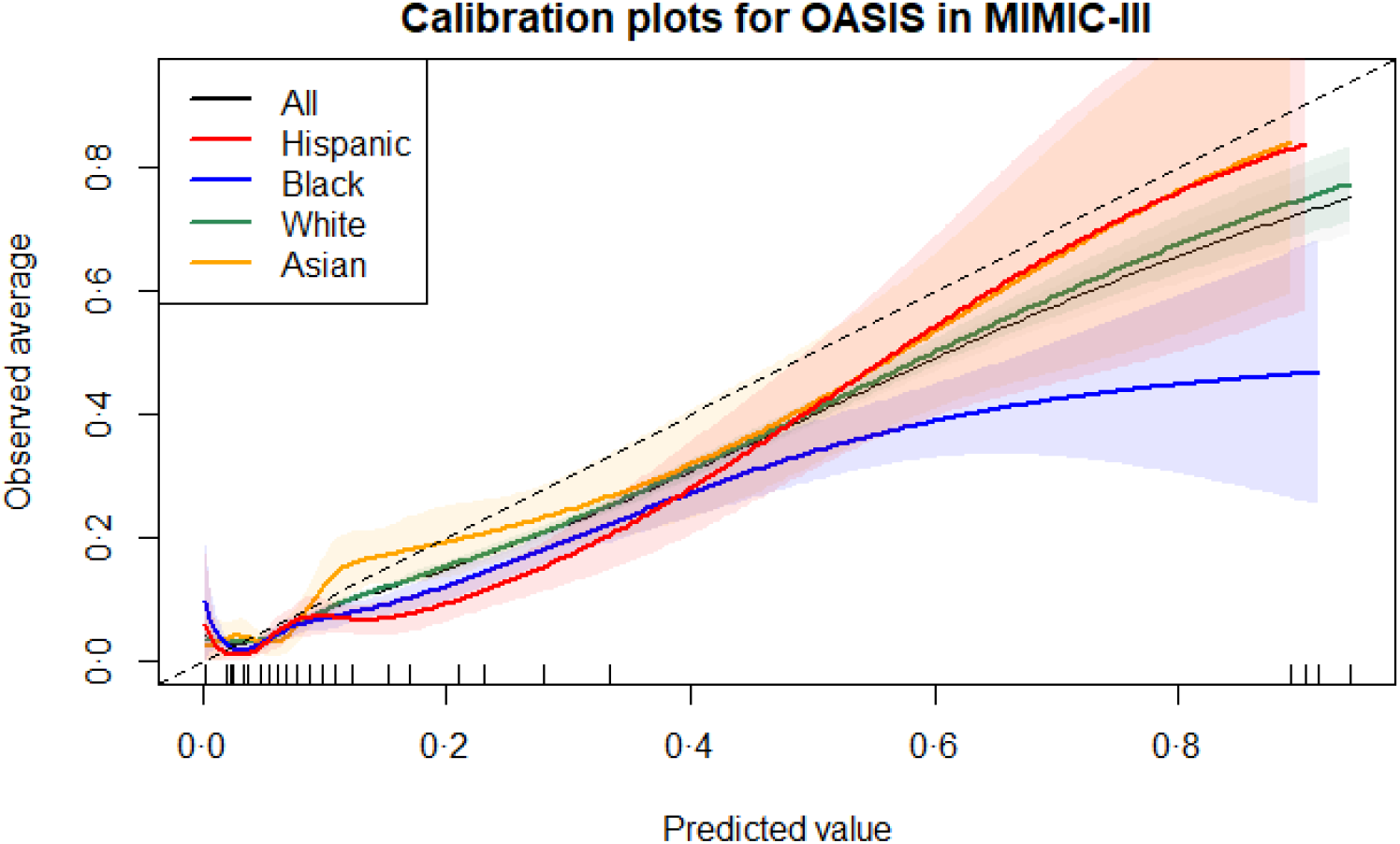

**Supplementary figure 8:**
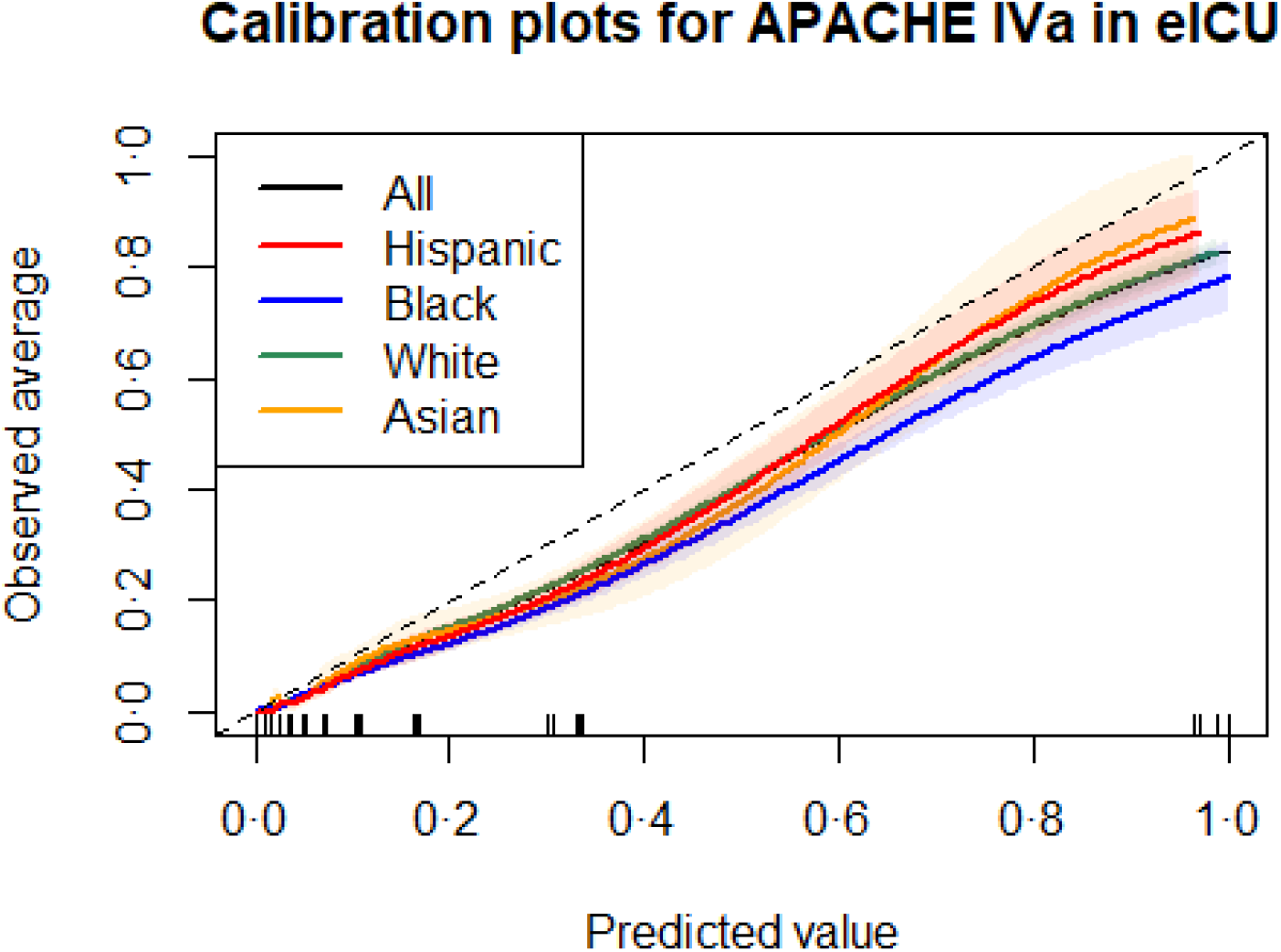

### Explanatory power of ethnicity and predictive scoring

**Supplementary table 2:** Logistic regression of APACHE IVa score and in-hospital death (eICU)

|  | Estimate | Std. Error | z value | Pr(>\|z\|) |
| --- | --- | --- | --- | --- |
| (Intercept) | -5.519 | 0.034 | -164.583 | 0 |
| apachescore | 0.047 | 0.000 | 114.491 | 0 |

$R^2 = 0.1972056$ .

**Supplementary table 3:** Logistic regression of OASIS score and in-hospital death(MIMIC-III)

|  | Estimate | Std. Error | z value | p-value |
| --- | --- | --- | --- | --- |
| (Intercept) | -5.890 | 0.071 | -83.531 | <0.0001 |
| OASIS | 0.111 | 0.002 | 60.054 | <0.0001 |

$R^2 = 0.1236887$

**Supplementary table 4:** Logistic regression of admission SOFA score and in-hospital death (MIMIC-III)

|  | Estimate | Std. Error | z value | p-value |
| --- | --- | --- | --- | --- |
| (Intercept) | -3.472 | 0.032 | -108.508 | <0.0001 |
| SOFA | 0.273 | 0.005 | 58.945 | <0.0001 |

$R^2 = 0.11774$

**Supplementary table 5:** Logistic regression of SOFA score and in-hospital death (eICU)

|  | Estimate | Std. Error | z value | Pr(>\|z\|) |
| --- | --- | --- | --- | --- |
| (Intercept) | -3.821 | 0.021 | -179.985 | 0 |
| SOFA1 | 0.313 | 0.003 | 97.837 | 0 |

$R^2 = 0.1177927$ .

**Supplementary table 6:** Logistic regression of ethnicity and in-hospital death (eICU), with African American as baseline ethnicity

|  | Estimate | Std. Error | z value | Pr(>\|z\|) |
| --- | --- | --- | --- | --- |
| (Intercept) | -2.447 | 0.030 | -81.951 | 0.000 |
| Asian | 0.138 | 0.086 | 1.607 | 0.108 |
| White | 0.092 | 0.032 | 2.896 | 0.004 |
| Hispanic | 0.101 | 0.058 | 1.739 | 0.082 |

$R^2 = 7.318419710^{-5}$ .

**Supplementary table 7:** Logistic regression of ethnicity and in-hospital death (MIMIC-III), with Asian as baseline ethnicity

|  | Estimate | Std. Error | z value | p-value |
| --- | --- | --- | --- | --- |
| (Intercept) | -1.890 | 0.086 | -22.007 | <0.0001 |
| African American | -0.408 | 0.099 | -4.105 | <0.0001 |
| Hispanic | -0.620 | 0.124 | -4.991 | <0.0001 |
| White | -0.157 | 0.087 | -1.798 | 0.072 |

$R^2 = 0.0011754$

**Supplementary table 8:** Logistic regression of ethnicity and in-hospital death controlled for APACHE IVa score (eICU), with African American as baseline ethnicity

|  | Estimate | Std. Error | z value | Pr(>\|z\|) |
| --- | --- | --- | --- | --- |
| (Intercept) | -5.689 | 0.048 | -119.610 | 0.000 |
| Asian | 0.208 | 0.098 | 2.125 | 0.034 |
| White | 0.186 | 0.036 | 5.109 | 0.000 |
| Hispanic | 0.184 | 0.066 | 2.778 | 0.005 |
| Apache score | 0.047 | 0.000 | 114.483 | 0.000 |

$R^2 = 0.1976472$ .

**Supplementary table 9:** Logistic regression of ethnicity and in-hospital death controlled for OASIS score <u>(MIMIC-II), with Asian as baseline ethnicity</u>

|  | Estimate | Std. Error | z value | p-value |
| --- | --- | --- | --- | --- |
| (Intercept) | -5.654 | 0.114 | -49.391 | <0.0001 |
| African American | -0.434 | 0.106 | -4.085 | <0.0001 |
| Hispanic | -0.500 | 0.132 | -3.802 | <0.0001 |
| White | -0.202 | 0.094 | -2.156 | 0.031 |
| OASIS | 0.111 | 0.002 | 59.904 | <0.0001 |

$R^2 = 0.1246951$

**Supplementary table 10:** Logistic regression of ethnicity and in-hospital death controlled for SOFA score (eICU), with African American as baseline ethnicity

|  | Estimate | Std. Error | z value | Pr(>\|z\|) |
| --- | --- | --- | --- | --- |
| (Intercept) | -4.005 | 0.037 | -107.052 | 0.000 |
| ethnicityAsian | 0.317 | 0.092 | 3.460 | 0.001 |
| ethnicityCaucasian | 0.205 | 0.034 | 6.013 | 0.000 |
| ethnicityHispanic | 0.168 | 0.062 | 2.711 | 0.007 |
| SOFA1 | 0.314 | 0.003 | 97.937 | 0.000 |

$R^2 = 0.1177927$ .

**Supplementary table 11:** Logistic regression of ethnicity and in-hospital death controlled for admission SOFA score (MIMIC-II), with Asian as baseline ethnicity

|  | Estimate | Std. Error | z value | p-value |
| --- | --- | --- | --- | --- |
| (Intercept) | -3.282 | 0.096 | -34.102 | <0.0001 |
| African American | -0.499 | 0.106 | -4.693 | <0.0001 |
| Hispanic | -0.638 | 0.133 | -4.801 | <0.0001 |
| White | -0.147 | 0.094 | -1.568 | 0.117 |
| SOFA | 0.274 | 0.005 | 59.050 | <0.0001 |

$R^2 = 0.119408$

### The contribution of the risk scores and ethnicity to variation in in-hospital mortality in the eICU-CRD and MIMIC-III databases

The Oasis score explains 12·367% of the variation in mortality, and the ethnicity explains 0·115% in MIMIC-III.

The APACHE IVa score explains 19·759% of the variation in mortality, and the ethnicity explains 0·055% in eICU-CRD

The SOFA score explains 11·837% of the variation in mortality, and the ethnicity explains 0·189% in the MIMIC-II database.

The SOFA score explains 11·801% of the variation in mortality, and the ethnicity explains 0·032% in the eICU-CRD database.

